# Feasibility and Validity of Ultra- Low-Field MRI for Measurement of Regional Infant Brain Volumes in Structures Associated with Antenatal Maternal Anemia

**DOI:** 10.1101/2025.06.10.25329262

**Authors:** Jessica E. Ringshaw, Niall J. Bourke, Michal R. Zieff, Catherine J. Wedderburn, Chiara Casella, Layla E. Bradford, Simone R. Williams, Donna Herr, Marlie Miles, Jonathan O’Muircheartaigh, Carly Bennallick, Khula South Africa Data Collection Team, Sean Deoni, Dan J. Stein, Daniel C. Alexander, Derek K. Jones, Steven C.R. Williams, Kirsten A. Donald

## Abstract

**Introduction:** The availability of ultra- low-field (ULF) magnetic resonance imaging (MRI) has the potential to improve neuroimaging accessibility in low-resource settings. However, the utility of ULF MRI in detecting child brain changes associated with anemia is unknown.

**Aim:** The aim of this study was to assess the comparability of 3T high-field (HF) and 64mT ULF volumes in infants for brain regions associated with antenatal maternal anemia.

**Method:** This neuroimaging sub-study is nested within Khula South Africa, a population-based birth cohort. Pregnant women were enrolled antenatally and postnatally, and mother-child dyads (*n* = 394) were followed prospectively at approximately 3, 6, 12 and 18 months. A sub-group of infants was scanned on 3T and 64mT MRI systems across study visits and images were segmented using MiniMORPH. Correlations and concordance coefficients were used to cross-validate HF and ULF infant brain volumes for the caudate nucleus, putamen, and corpus callosum.

**Results:** 78 children (53.85% male) had paired HF (*Mean* [*SD*] age = 9.64 [5.26] months) and ULF (*Mean* [*SD*] age = 9.47 [5.32] months) datasets. Results indicated strong agreement between systems for intracranial volume (ICV; *r* = 0.96, ρ_ccc_ = 0.95), and brain regions of interest in anemia including the caudate (*r* = 0.89, ρ_ccc_ = 0.86), putamen (*r* = 0.97, ρ_ccc_ = 0.96), and corpus callosum (*r* = 0.87, ρ_ccc_ = 0.79).

**Conclusion:** This cross-validation study demonstrates excellent correspondence between 3T and 64mT volumes for infant brain regions implicated in antenatal maternal anemia. Findings validate the use of ULF MRI for paediatric neuroimaging on anemia in Africa.

**HIGHLIGHTS:**

- This cross-validation study is the first to compare HF and ULF volume estimates for infant brain regions previously found to be associated with antenatal maternal anemia within the first two years of life.
- Key findings of this research demonstrate linear associations and strong agreement between HF and ULF volume estimates for the caudate nucleus, putamen, and corpus callosum in infants between 3-18 months of age. Improved correspondence between HF and ULF MRI was observed in older infants, particularly for basal ganglia structures.
- These novel findings validate the use of ULF MRI for paediatric neuroimaging work on antenatal maternal anemia and other prevalent health priorities in low- and middle-income countries.

**GRAPHICAL ABSTRACT:** This cross-validation study assessed the comparability of high-field (3T) and ultra- low-field (64mT) volumes in infants for brain regions associated with antenatal maternal anemia. Key findings demonstrated strong agreement between HF and ULF volume estimates for the caudate nucleus, putamen, and corpus callosum in infants between 3-18 months of age.

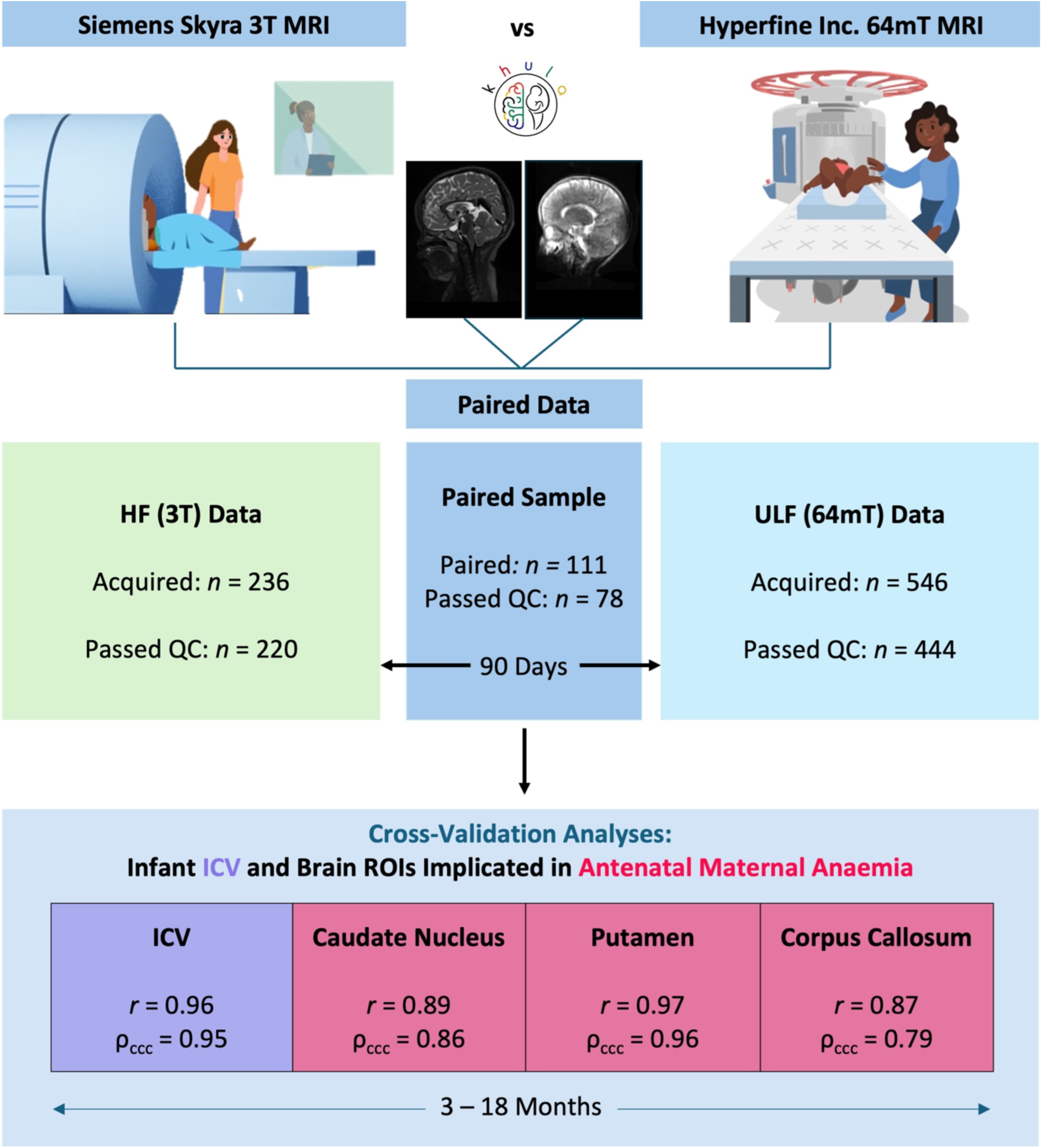

## INTRODUCTION

Anemia, a highly prevalent condition characterised by low serum hemoglobin, is a global health concern affecting approximately 571 million women.^1,2^ While all women of reproductive age are at risk of anemia, this risk is exacerbated during pregnancy when biological demands for the hemoglobin-facilitated delivery of oxygenated blood increases to support the growing foetus.^3,4^ Hemoglobin production relies on iron and is vital for the healthy maturation of the fetal brain in utero as well as the process of fetal iron loading prior to birth.^5^ While there is a well-established association between antenatal maternal anemia and poor cognitive outcomes in children,^6,7^ including evidence from South Africa,^8^ less is known about the timing and precise underlying neurobiological mechanisms. An improved understanding of this relationship may inform the timing and optimisation of prevention and intervention strategies for anemia in pregnancy and infancy.^9^

Neuroimaging tools such as Magnetic Resonance Imaging (MRI) bridge this gap by providing a non-invasive and objective measure of assessing brain structure.^10^ A few studies, using high-field (HF) MRI, have suggested that the developing brain may be particularly sensitive to the effects of antenatal maternal anemia.^11–13^ This is reinforced by findings from a birth cohort in South Africa suggesting that these effects may be regionally consistent and tend to persist with age in children from 2-3 years^9^ through to age 6-7 years.^14^ This highlights a need for corroboratory research in other high-risk settings and across timepoints, including the first two years of life.

Traditional HF MRI systems (≥1.5T) are expensive, requiring significant infrastructure, specialised expertise, and a high power supply.^15,16^ This technology has limited accessibility in most low- and middle-income countries (LMICs) with an average of only 0.2-1.1 MRI scanners per million people compared to high-income countries where there are approximately 26.53 MRI scanners per million people.^17^ As a result, there is disproportionately less neuroimaging research emerging from LMICs, particularly in sub-Saharan Africa, despite the burden of maternal anemia being striking in this region with 30-50% of pregnant women estimated to be anemic.^1^ In an effort to address this neuroimaging disparity, there has been an increased focus on developing scalable tools and methods that are practical for use in under-resourced settings. This includes validating the use of a novel ultra-low-field (ULF) MRI system (Hyperfine, Inc. 64mT).^18^ This mobile scanner is more cost-effective, requires less power, and is less noisy than conventional systems, allowing it to be easily integrated into low-resource settings for paediatric use.^19^

Thus far, a preliminary validation study using the ULF 64mT system on neurotypical children between 6 weeks and 16 years of age in high-income settings has been positive, demonstrating strong associations between volumes extracted from structural sequences on paired HF and ULF MRI.^20^ Similarly, in a study with both HF (3T) and ULF (64mT) scans, structural sequences were found to be useful for neuroanatomical identification and the detection of discrete brain abnormalities in a clinical sample of neonates in the Intensive Care Unit (ICU).^21^ However, the utility of these systems in detecting group differences in regional brain structures associated with known risk factors of varying effect sizes for neurodevelopmental impact is unknown. Therefore, scanners have been implemented in clinical research sites across sub-Saharan Africa and South Asia as a complementary tool in ongoing research on prevalent and context-specific health priorities such as malnutrition and anemia.^22^

In order to meaningfully interpret any neuroimaging findings for this important clinical research, it is necessary to determine the comparative utility of the ULF 64mT system for estimating brain volumes relative to HF MRI, with 3T typically being considered as the conventional gold standard. Cross-validation work of this nature has recently demonstrated excellent correspondence between structural outputs on 3T and 64mT MRI in healthy adults for four global tissue types and 98 local structures, with stronger concordance in larger brain regions.^23^ While promising, these conclusions were based on data from a sample size of 23 healthy adults. Correspondence is likely to be lower in paediatric samples due to both increased challenges acquiring data (including risks of motion artefact, poorer contrast due to incomplete myelination, and a higher water content in the developing brain), and segmenting data (segmentation inaccuracies associated with smaller absolute size of structures and the limited availability of processing pipelines with age-appropriate templates and atlases for segmentation of infant and child data).^24–28^ However, many of these issues are being mitigated by the implementation of clinical strategies for acquiring high quality data by successfully scanning infants in natural sleep,^29^ developments in ULF imaging hardware and software,^22^ and the optimisation of processing pipelines for paediatric data.^22,30–32^ In order to assess the current feasibility of ULF MRI, similar cross-validation work needs to be conducted using paired HF and ULF data from a paediatric sample with a specific focus on brain regions affected by prevalent risk factors of interest such as anemia.

We conducted this research in infants (aged 3-18 months) who were scanned on both HF and ULF MRI scanners as part of the Khula study^33^ in South Africa. Recent epidemiological research from this cohort revealed that approximately one third of pregnant mothers were anemic and half were iron deficient after adjustment for inflammation, with 50% of antenatal anemia cases attributable to iron deficiency.^34^ Given previously demonstrated associations between antenatal maternal anemia and child brain structure using HF MRI, there is a need to further investigate such relationships in infants. While many other high-risk settings do not have access to paired HF and ULF MRI, the Khula study offers the opportunity to inform the feasibility of ULF MRI for research on anemia, particularly in LMICs. The aim of this study was to compare HF and ULF volume estimates for infant brain regions previously implicated with antenatal maternal anemia within the first two years of life.

## METHODS

### Study design and setting

This neuroimaging sub-study is nested within the Khula^33^ study in South Africa, an observational population-based birth cohort with a multi-modal approach to investigating risk and protective factors for paediatric neurodevelopment. Mothers were recruited during the third trimester of pregnancy from antenatal clinic visits at the Gugulethu Midwife Obstetrics Unit (MOU; primary healthcare centre) and in the early postnatal period from nearby clinics. Gugulethu is an urban informal settlement at sea level in the Western Cape, approximately 18km from Cape Town, with predominantly isiXhosa speaking residents. The community is characterised by a high prevalence of psychosocial and health exposures including interpersonal violence, maternal depression, malnutrition, and infection including Human Immunodeficiency Virus (HIV). All mother-child dyads enrolled in Khula were followed prospectively and attended study visits at approximately 3 months, 6 months, 12 months, and 18 months. A sub-group of infants were scanned on Siemens 3T Skyra (HF) and Hyperfine, Inc. 64mT scanner (ULF; software versions 8.2–8.6.1) systems within 90 days of each other across study visits.

### Participants

Eligibility for recruitment (December 2021 – November 2022) of mothers required being over the age of 18 years and in the third trimester of pregnancy (28-36 weeks) or up to 3-6 months after childbirth. Exclusion criteria included multiple pregnancies, psychotropic drug use during pregnancy, infant congenital malformations or abnormalities (e.g. Spina Bifida, Down’s Syndrome), and severe birth complications (e.g. uterine rupture, birth asphyxia).

For the umbrella study, 394 mother-child pairs were enrolled in Khula, with 329 mothers recruited antenatally and 65 mothers recruited postnatally. Of these dyads, 236 HF scans and 546 ULF scans were collected across study visits between 3 and 18 months. All scans were subject to a rigorous visual quality check (QC) procedure performed by a senior neuroimaging staff member using a standardised protocol that assesses image quality based on observable indications such as motion artefact, contrast, electrical interference, signal-to-noise ratio, and field-of-view. This procedure was followed using a step-by-step form on the Flywheel platform, as per study data management protocol.^22^ Any scans that failed QC were excluded from analyses, leaving 220/236 useable HF scans (93.22% success rate) and 444/546 useable ULF scans (81.32% success rate). The paired dataset (*n* = 78) was compiled for a pooled sample of infants across age groups with both useable HF and ULF scans acquired within 90 days of one another and processed with the same age template (see Neuroimaging section below). The paired sample determination is illustrated in the data flowchart (Figure 1).

**Figure 1.**
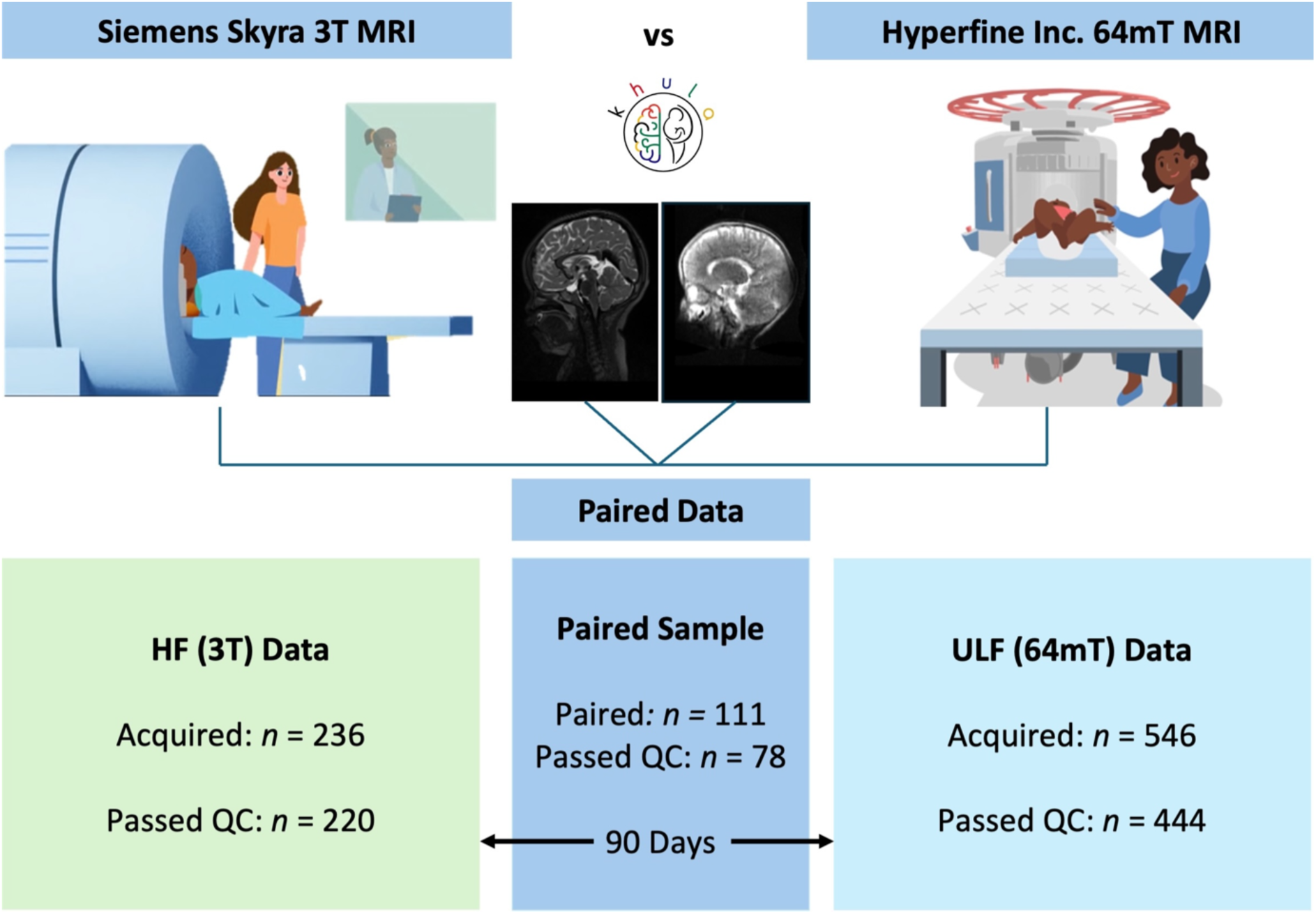
Study Flowchart Demonstrating the Determination of a Paired Sample with Infant Scans from HF (3T) and ULF (64mT) MRI Systems. *Legend.* Animated images were created for neuroimaging explainer videos directed by J.E. Ringshaw as a public engagement project linked to the Khula study in South Africa. Available online for HF^35,36^ and ULF^37^ scanning protocols.

### Measures

#### Demographics

Child sex and age at the time of scan was collected. Individual infant datasets (HF and ULF scans) were paired for this analysis if their age at the time of scan acquisition fell within the same age group used for template registration during processing. Additional demographic data on socio-economic status (SES; indicated by maternal education and household income), maternal employment, and antenatal maternal anemia status was reported to provide context for the sample.

#### Neuroimaging

##### Scan acquisition

Both HF (3T; Siemens Skyra) and ULF (64mT; Hyperfine Inc.,) imaging was conducted at the Cape Universities Body Imaging Centre (CUBIC). Neuroimaging data were collected during natural sleep using strategies developed and used successfully for paediatric scanning by our team.^29^ These include adequate preparation (desensitization, screening, rapport-building) incorporating the use of locally developed neuroimaging explainer videos for <u>HF</u>^35,36^ and <u>ULF</u>^37^ scanning protocols, baby-friendly rooms, strategic scan timing (after other data collection activities when babies are most tired and coinciding with naps in older infants), breastfeeding (and subsequently burping to avoid discomfort due to gas) or warm meals and nappy changes prior to sleep initiation, context-specific sleep positioning (typically babies are swaddled on their mother’s backs in South Africa), consistency in background noise, adequate ventilation and comfortable room temperatures, noise protection (ear plugs), immobilizing cushions for securing the head, and close body contact between research staff and babies during transfer and positioning in the scanner.

For HF MRI, T2-weighted sequences were acquired on a 3T Siemens Skyra system in the sagittal orientation using 16-channel (for infants at 3- and 6-month visits) and 32-channel (for infants at 12- and 18-month visits) head coils with the following parameters: repetition time=3200ms; echo time= 561ms; voxel size 1.0x1.0x1.0mm^3^; field of view=256mm x 256mm; 144 slices, 1.0mm thick. Scan time: 3min30s. ULF MRI scans were acquired on the <u>Hyperfine Inc.,</u> 64mT system (software versions 8.2–8.6.1). Given that T2-weighted sequences are currently the most developed at ULF, and best for structural imaging in infancy, they were chosen for comparative purposes. Each plane was acquired separately as orthogonal anisotropic images for axial (1.5mm x 1.5mm x 5mm), coronal (1.5mm x 5mm x 1.5mm), and sagittal (5mm x 1.5mm x 1.5mm) sequences. All HF scans were reviewed and reported on by a clinical radiologist. Any qualitative abnormalities or incidental findings were discussed with a paediatric neurologist and, where appropriate, referred via established clinical pathways. For this study, no incidental findings were observed and, therefore, no scans were excluded from the analyses on this basis.

##### Neuroimaging processing pipelines

After quality checking both HF and ULF raw data using the standardized protocol described above (Participants section), only useable scans were included in analyses. Based on previous HF findings on antenatal maternal anemia,^9,14^ volumes for the corpus callosum, caudate nucleus, and putamen were chosen as regions of interest (ROIs).

Using a multi-resolution registration (MRR) technique, ULF T2W scans were reconstructed from three orthogonally-acquired anisotropic images (axial, coronal, sagittal) into a single 1.5mm^3^ isotropic images of higher effective resolution.^38^ Thereafter, images across field strengths (64mT and 3T) were segmented using <u>MiniMORPH</u>^32^ (Figure 2). Tissue and cerebrospinal fluid (CSF) priors were generated by aligning Baby Connectome Project (BCP)^39^ probability maps to native isotropic images via age-specific templates^40^ derived from Khula study data. These priors and probability maps were used to segment T2w images in ANTs.^41^ Subcortical and callosal parcellations from BCP^39^ and Penn-CHOP^42^ atlases were also registered to native isotropic images.^40^ The corpus callosum was manually segmented based on the Desikan-Killiany atlas.^43^ The tissue class obtained from ANTs was multiplied by subcortical grey matter and callosal masks to refine subcortical and callosal segmentations.

**Figure 2.**
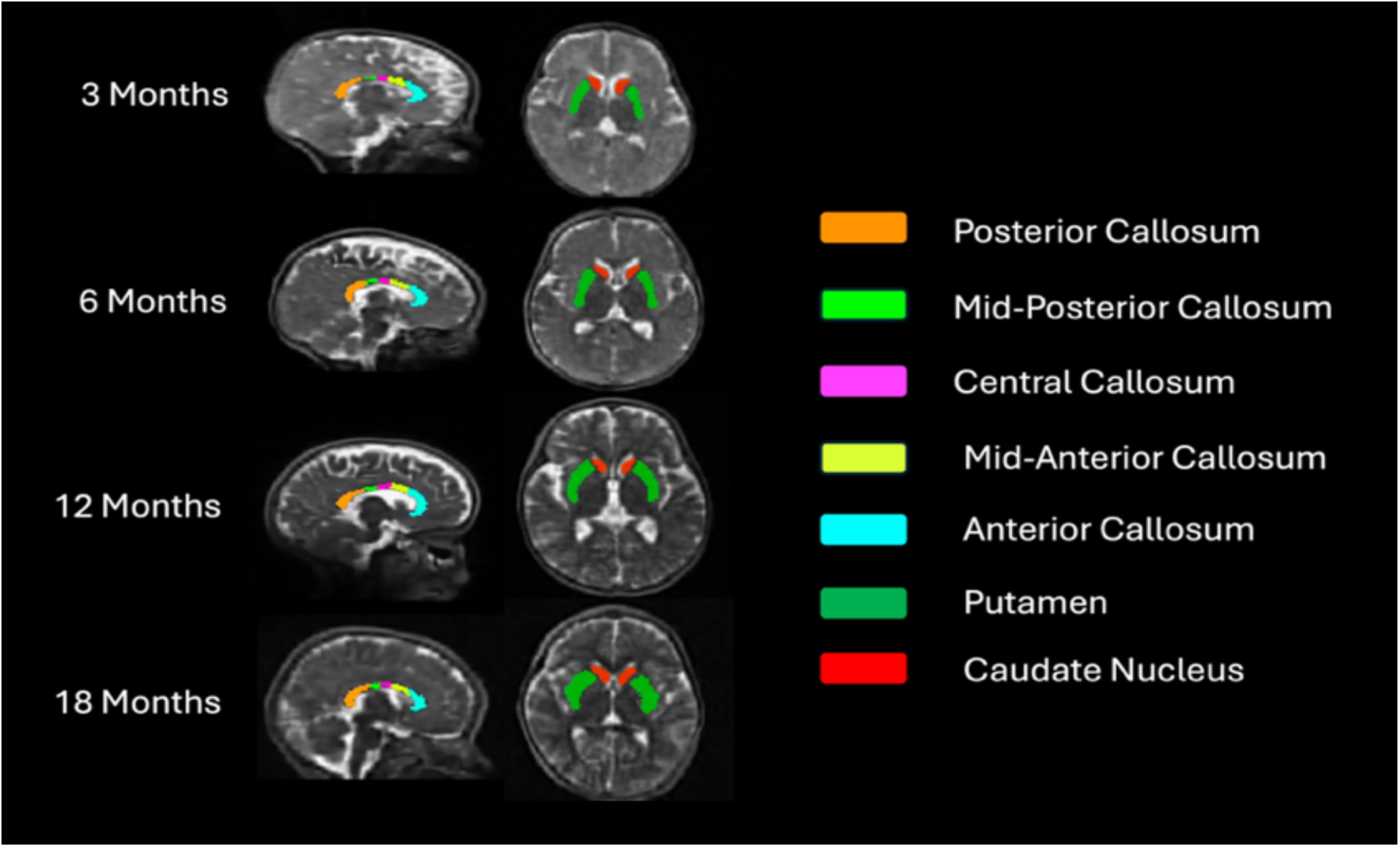
ULF (64mT) MRI Data Across Imaging Age Groups: Examples of Segmented ROIs Overlaid on Sagittal and Axial Views of T2W Isotropic Images.

The previously developed MRR^38^ and MiniMORPH^32^ processing pipelines were integrated to run using an automated gear implemented on Flywheel for the batch-processing all scans. The MiniMORPH gear was set to run based on the child’s age at the time of scan, using age-specific templates for 3 months (scan age < 5 months), 6 months (scan age <10 months), 12 months (scan age < 16 months), and 18 months (scan age < 22 months).

### Statistical Analysis

Given the broad age windows of the study timepoints, individual infant datasets (HF and ULF scans) were paired based on their age at the time of scan acquisition and the corresponding age group selected for registration using Khula age-specific templates. A Z-score criterion of > 3 or < -3 was used to identify outliers for regional infant brain volumes assessed separately for HF and ULF data across each respective age group. Given that no outliers were observed, no paired scans were excluded from the sample on this basis. The characteristics of the pooled sample and each age group were presented as means and standard deviations for continuous variables and frequencies for categorical variables. To test for group differences in age between scans acquired for the same children at HF and ULF, paired t-tests were conducted. All statistical analyses were conducted in R and a two-sided significance level was used throughout (*p* < 0.05).

The comparative relationship between HF and ULF MRI was assessed using Pearson’s correlation coefficients (*r*) and Lin’s concordance correlation coefficients (ρ_ccc_) in a pooled sample of paired scans. While the correlation coefficients quantified the strength and direction of the relationship between HF and ULF volumes, the concordance coefficients demonstrated the level of agreement between HF and ULF volume estimates. This statistical analysis approach was repeated for each age group separately to assess the comparative performance and feasibility of ULF MRI in younger versus older children.

In the pooled sample with an age-specific key, the associations between HF and ULF volume estimates were graphically represented using a regression line of best fit (Figure 3). This was plotted relative to an identity line demonstrating perfect concordance (where HF and ULF are equivalent; *Y = X*) visualized as a 45° angle on axes with the same scale and dynamic range. The regression equation was used to quantify the magnitude and direction of any disagreement between HF and ULF volume estimates. Specifically, the slope (β₁) of the regression was used to assess how ULF volumes predicted HF volumes, assuming HF as the reference. Given an intercept (β_0_) of 0, a slope < 1 indicated systematic overestimation of HF volumes by ULF volumes (i.e., ULF values are proportionally larger than HF), while a slope > 1 indicated systematic underestimation. However, in parallel, the intercept (value of *Y* when *X* = 0) and the crossover point (where the regression line intersects the line of identity, *Y = X*) were used to identify the value of *X* at which the direction of bias may switch between overestimation and underestimation. Furthermore, a General Linear Hypothesis Test (GLHT) was used to jointly determine whether the regression model was significantly different from the line of identity (*Y = X*), as indicated by a slope that differed significantly from 1 and an intercept that differed significantly from 0. This assumed that a slope of 1 and an intercept of 0 represents perfect agreement. For brain regions with significant disagreement, the extent to which the ULF overestimated or underestimated HF regional brain volumes was quantified using percentage differences for each age group.

**Figure 3.**
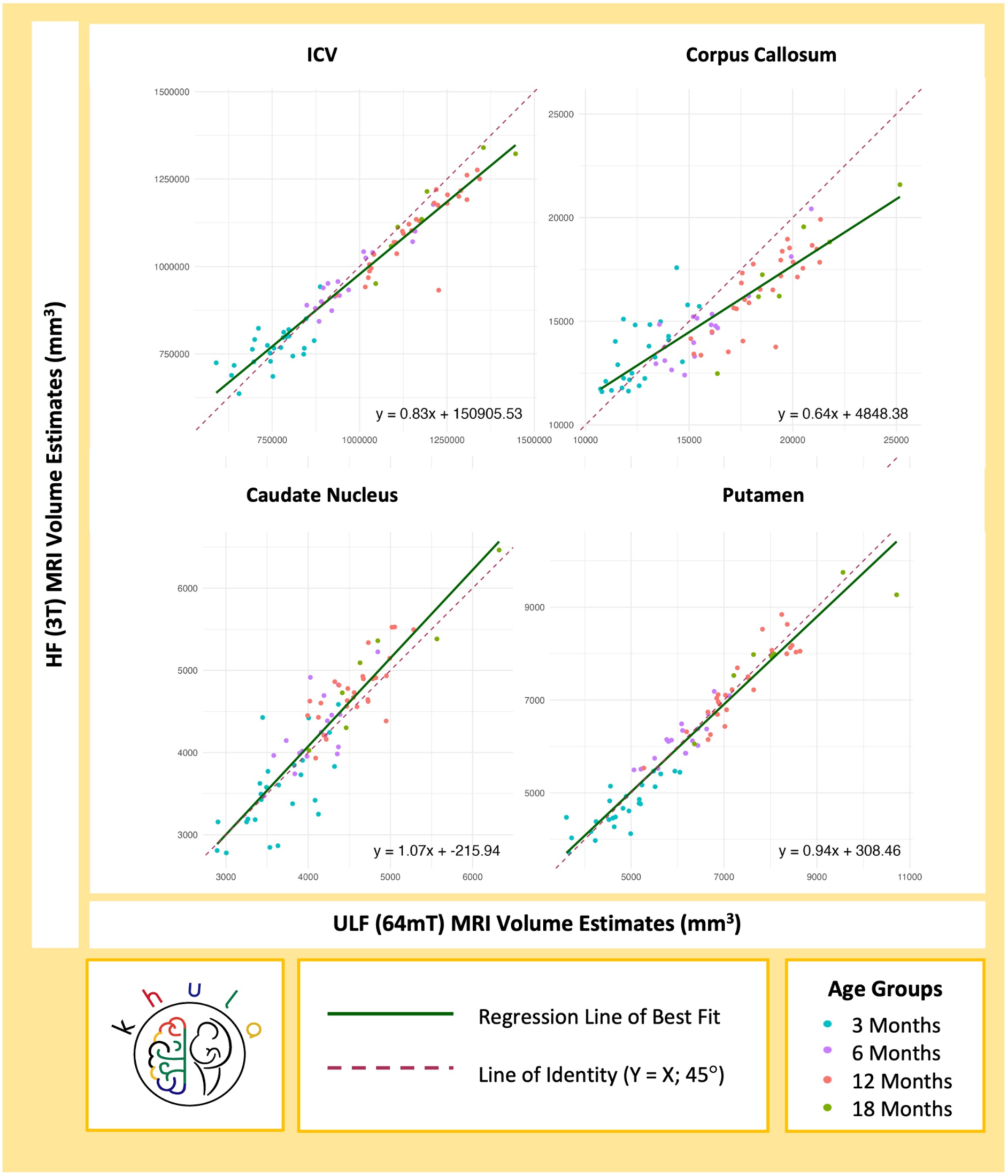
Associations and Concordance Between HF (3T) and ULF (64mT) MRI Volume Estimates for ICV and Regional Brain Volumes (mm^3^) Implicated in Anemia.

Lastly, agreement between HF and ULF systems for ICV and regional brain volumes was explored further using Bland-Altman plots (Figure 4). The difference in HF and ULF volumes was plotted against the average brain volume measured by the two systems, with a key for age group. The mean difference line demonstrated the average bias between HF and ULF MRI, with readings closer to 0 representing stronger agreement. The limits of agreement displayed the expected range within which 95% of differences occurred, with narrow limits indicating less variability.

**Figure 4.**
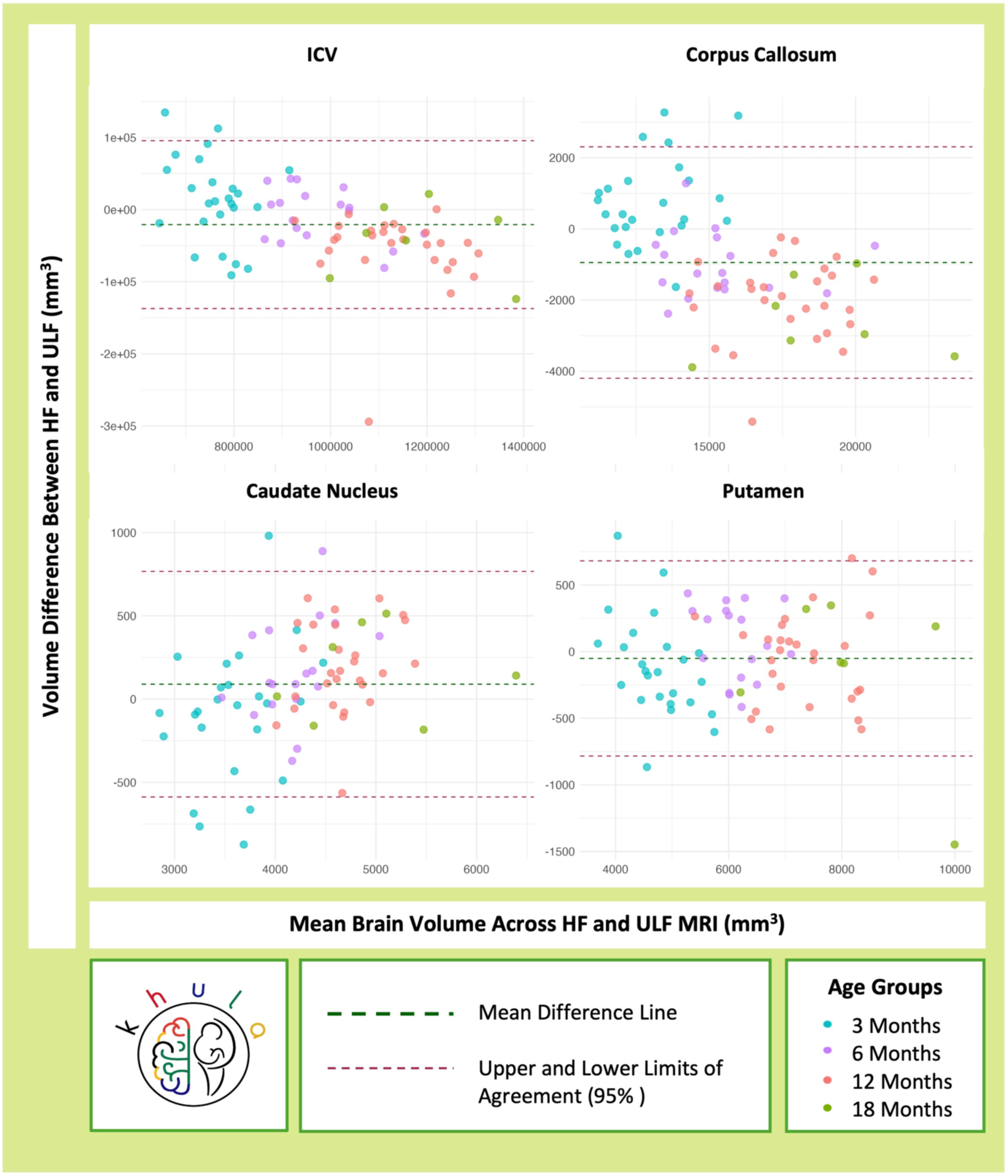
Bland-Altman Plots Demonstrating the Difference Between HF (3T) and ULF (64mT) Volume Estimates for the Corpus Callosum Volume Across Age Groups.

## RESULTS

### Sample Characteristics

Across all study timepoints, 78 children (42 [53.85%] male) had paired HF (*Mean*[*SD*] age = 9.64 [5.26] months) and ULF (*Mean* [*SD*] = 9.47 [5.32] months) datasets. In this sample, 47.44% (41/78) of mothers had not completed secondary school, 64.10% (50/78) of mothers were unemployed, and 66.67% (52/78) of households had a combined income of less than R5000 (approximately $300) per month. Overall, 65 mothers had antenatal maternal hemoglobin data, of which 26.15% (17/65) were found to be anemic during pregnancy.

There were no differences in age between children with HF and ULF for paired groups at 3 months, 6 months, and 18 months (Table 1). However, in the 12-month age group, children were found to be significantly older at the acquisition of their HF scan (*Mean* [*SD*] = 13.60 [1.91] months than their ULF scan (*Mean* [*SD*] = 13.15 [1.76] months), *p* = 0.032.

**Table 1.** Age Distribution Across HF and ULF MRI Acquisitions in a Paired Sample (*n* = 78)

| Khula Study Visit Number | Age Group in Months<sup>a</sup> | Mean Age (Months)<br>M (SD) [Range] | | $p^b$ |
| --- | --- | --- | --- | --- |
|  |  | HF (3T)<br>MRI Scan | ULF (64mT)<br>MRI Scan |  |
| 1 | 3 ($n = 25$) | 3.72 (0.68)<br>[2.56 – 5.00] | 3.58 (0.93)<br>[2.27 – 4.96] | 0.405 |
| 2 | 6 ($n = 18$) | 8.10 (1.37)<br>[5.39 – 9.96] | 7.90 (1.07)<br>[5.39 – 9.60] | 0.463 |
| 3 | 12 ($n = 28$) | 13.60 (1.91)<br>[10.42 – 15.81] | 13.15 (1.76)<br>[9.93 – 15.39] | 0.032 |
| 4 | 18 ($n = 7$) | 18.94 (2.15)<br>[16.54 – 21.07] | 19.78 (1.79)<br>[16.77 – 21.90] | 0.140 |
| Across Visits | 3 – 18 ($n = 78$) | 9.64 (5.26)<br>[2.56 – 21.07] | 9.47 (5.32)<br>[2.27 – 21.90] | 0.151 |

<sup>a</sup> Age groups correspond to the age-specific templates used for processing scans with MiniMORPH. The age-specific template was selected according to the infants actual age at the time of the scan. HF and ULF MRI scans were paired if they had been processed with the same age-specific template.
<sup>b</sup> A paired t-test was conducted to assess differences in age between HF MRI and ULF MRI acquisitions within the same sample.

### Cross-Validation: Full Sample

In the pooled sample with all age groups, volume estimates were significantly correlated (*p* < 0.001) with a high degree of concordance for total intracranial volume (ICV; *r* = 0.96, ρ_ccc_ = 0.95), and brain ROIs in antenatal maternal anemia including the putamen (*r* = 0.97, ρ_ccc_ = 0.96), caudate nucleus (*r* = 0.89, ρ_ccc_ = 0.86), and corpus callosum (*r* = 0.87, ρ_ccc_ = 0.79). Across all age groups, the putamen emerged as having the strongest comparative relationship between HF and ULF volume estimates. Overall, Pearson’s correlations and Lin’s concordance coefficients were similar, demonstrating a linear relationship with strong agreement between brain volume estimates in infants between 3-18 months of age (Figure 3).

Using a joint GLHT, the linear regression model (Figure 3) for the putamen (Y = 0.94*X* + 308.46) did not differ significantly from the line of identity, demonstrating theoretical agreement (slope = 1) without bias (intercept = 0), *F* (2,76) = 2.68, *p* = 0.075. While there was strong concordance between HF and ULF volumes for the caudate nucleus, it did not represent perfect agreement as the regression line (*Y* = 1.07*X* – 215.94) differed slightly from the line of identity, *F* (2,76) = 3.27, *p* = 0.043. While the slope was marginally > 1 suggesting systematic underestimation, the negative intercept was indicative of a downward bias and there was crossover between the regression and identity lines (Figure 3). Taken together, this model predicted that ULF overestimated HF MRI at lower brain volume ranges (younger age groups) and underestimated them at higher brain volume ranges (older age groups). The disagreement between HF and ULF MRI was more prominent for the corpus callosum (*Y* = 0.64*X* + 4848.38; F [2,76] = 62.69, p < 0.001) and ICV (*Y* = 0.83*X* + 150905.53; F(2,76) = 29.40, p<0.001) with the regression models differing to a greater extent from the identity lines for both. In these instances, regression slopes of <1 with a positive intercept and some crossover (Figure 3) suggested ULF underestimation of HF at lower brain volumes (younger age groups) and overestimation at higher brain volumes (older age groups). The nature and magnitude of disagreement was explored further by age groups below.

### Cross-Validation: By Age Group

Cross-validation analyses were repeated for each individual age group to assess the relative predictive performance of ULF MRI in younger and older children (Table 2). These findings are discussed separately for each brain region below.

**Table 2.** Pearson’s Correlations and Concordance Coefficients for ICV and Brain ROIs in Antenatal Maternal Anemia for Infants Across Age Groups (*n* = 78)

| Brain Region | Correlation and Concordance ($r$, $\rho_{ccc}$)<sup>a</sup> | | | |
| --- | --- | --- | --- | --- |
| | Study Visit 1:<br>3 Months<sup>b</sup><br>($n = 25$) | Study Visit 2:<br>6 Months<sup>b</sup><br>($n = 18$) | Study Visit 3:<br>12 Months<sup>b</sup><br>($n = 28$) | Study Visit 4:<br>18 Months<sup>b</sup><br>($n = 7$) |
| Caudate Nucleus | 0.63*** (0.61) | 0.73*** (0.64) | 0.74*** (0.65) | 0.94** (0.92) |
| Putamen | 0.83*** (0.78) | 0.85*** (0.84) | 0.91*** (0.91) | 0.91** (0.89) |
| Corpus Callosum | 0.69*** (0.60) | 0.90*** (0.79) | 0.82*** (0.51) | 0.92** (0.63) |
| ICV | 0.64*** (0.61) | 0.94*** (0.93) | 0.88*** (0.78) | 0.93** (0.89) |

<sup>a</sup> The comparative relationship between HF and ULF MRI was assessed using Pearson's correlation coefficient ( $r$ ) and Lin's concordance correlation coefficients ( $\rho_{ccc}$ ) in a pooled sample of paired scans.
<sup>b</sup> Age groups correspond to the age-specific templates used for processing scans with MiniMORPH. The age-specific template was selected according to the infants actual age at the time of the scan. HF and ULF MRI scans were paired if they had been processed with the same age-specific template.
\*\* $p$ is significant at $<0.01$ , \*\*\* $p$ is significant at $<0.001$

#### ICV

Agreement between HF and ULF estimates (Table 2) for total brain volume, as indicated by ICV, strengthened with age from 3 months (*r* = 0.64, ρ_ccc_ = 0.61) to 18 months (*r* = 0.93, ρ_ccc_ = 0.89). However, there was an inconsistency at 12 months, with correlations and concordance dropping for this age group. Following on from the GLHT findings for the full sample (Figure 3), ULF underestimated HF estimates of ICV by 1.77% at 3 months, and overestimated them by 0.78% at 6 months, 4.97% at 12 months, and 3.48% at 18 months. This result was further supported by the Bland-Altman plot (Figure 4) which demonstrated a mean difference line (HF – ULF) just below 0 with slightly higher HF ICV volumes (> difference; underestimation) at 3 months and slightly lower HF volumes (< difference; overestimation) at 12 and 18 months.

#### Basal ganglia

The comparative association between HF and ULF volume estimates consistently improved with age for the putamen and the caudate nucleus (Table 2). By 18 months, both structures had strong positive correlations above 0.9 with similar concordance coefficients. As demonstrated using GLHT results for the full sample, HF and ULF volume estimates for the putamen were in agreement (Figure 3; Figure 4). Following on from the GLHT findings demonstrating marginal disagreement for the caudate nucleus in the full sample (Figure 3), ULF overestimated HF volumes for this structure by 2.60% at 3 months, and underestimated them by 3.79% at 6 months, 3.96% at 12 months, and 3.12% at 18 months. This is in alignment with the Bland-Altman plot (Figure 4), demonstrating a mean difference line (HF – ULF) just above 0, with slight overestimation (< difference) in infants at 3 months and slight underestimation (> difference) in infants at 6-18 months. However, this was marginal, with 95% of differences occurring within narrow limits of ±500 mm³.

#### Corpus callosum

There was an overall improvement (Table 2) in correlations and concordance coefficients with age for the corpus callosum from 3 (*r* = 0.69, ρ_ccc_ = 0.60) months to 18 months (*r* = 0.92, ρ_ccc_ = 0.63). However, there were inconsistencies across age groups with the strongest agreement for infants in the 6-month age group (*r* = 0.90, ρ_ccc_ = 0.79) followed by the weakest agreement for infants in the 12-month age group (*r* = 0.82, ρ_ccc_ = 0.51). Following on from the GLHT findings demonstrating disagreement and crossover in the full sample (Figure 3), it was found that the ULF system underestimated corpus callous volumes by 5.57% in children at 3 months, and overestimated them by 6.77% at 6 months, 12.15% at 12 months, and 14.72% at 18 months. This was corroborated by the Bland-Altman plot (Figure 4) which revealed a pattern of differences (HF – ULF) indicative of ULF underestimating HF corpus callous volumes in younger infants at 3 months and overestimating them in older infants at 6 – 18 months.

Overall, the magnitude of the % volume difference between HF and ULF volumes was largest in children at 12 and 18 months, demonstrating that overestimation in older age groups occurred to a greater extent than underestimation in the younger children. This was reinforced by the Bland-Altman plot (Figure 4) with a wider lower limit (-4000 mm³) showing negative differences (overestimation) in older infants than the upper limit (2000 mm³) showing positive differences (underestimation) in younger children.

To gain further insight into any misalignment between HF and ULF volume estimates for the corpus callosum, each segment was compared separately (Table 3). Overall, the concordance between HF and ULF volume estimates for the corpus callosum regions improved across the 3 and 6-month age groups. However, there was less agreement in the older children, particularly in the 12-month age group. While this age-related pattern was true for all corpus callosum regions, the mid-posterior region of the corpus callosum consistently demonstrated weaker associations and lower concordance coefficients across age groups than other corpus callosum regions (Table 3).

**Table 3.**
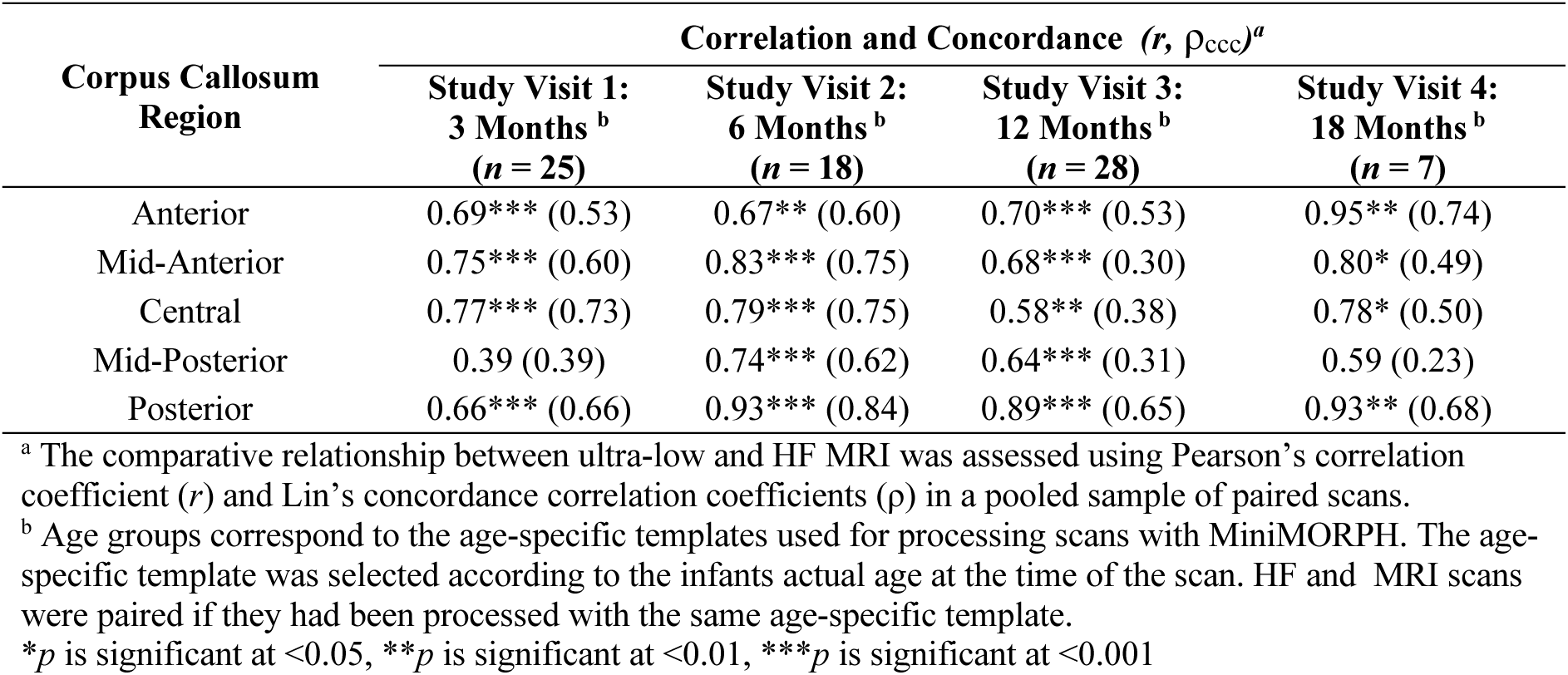
Pearson’s Correlations and Concordance Coefficients for Corpus Callosum Regions (*n* = 78)

| Corpus Callosum Region | Correlation and Concordance (*r*, $\rho_{ccc}$)<sup>a</sup> | | | |
| --- | --- | --- | --- | --- |
|  | Study Visit 1:<br>3 Months <sup>b</sup><br>(*n* = 25) | Study Visit 2:<br>6 Months <sup>b</sup><br>(*n* = 18) | Study Visit 3:<br>12 Months <sup>b</sup><br>(*n* = 28) | Study Visit 4:<br>18 Months <sup>b</sup><br>(*n* = 7) |
| Anterior | 0.69*** (0.53) | 0.67** (0.60) | 0.70*** (0.53) | 0.95** (0.74) |
| Mid-Anterior | 0.75*** (0.60) | 0.83*** (0.75) | 0.68*** (0.30) | 0.80* (0.49) |
| Central | 0.77*** (0.73) | 0.79*** (0.75) | 0.58** (0.38) | 0.78* (0.50) |
| Mid-Posterior | 0.39 (0.39) | 0.74*** (0.62) | 0.64*** (0.31) | 0.59 (0.23) |
| Posterior | 0.66*** (0.66) | 0.93*** (0.84) | 0.89*** (0.65) | 0.93** (0.68) |

<sup>a</sup> The comparative relationship between ultra-low and HF MRI was assessed using Pearson's correlation coefficient (*r*) and Lin's concordance correlation coefficients ( $\rho$ ) in a pooled sample of paired scans.
<sup>b</sup> Age groups correspond to the age-specific templates used for processing scans with MiniMORPH. The age-specific template was selected according to the infants actual age at the time of the scan. HF and MRI scans were paired if they had been processed with the same age-specific template.
\**p* is significant at <0.05, \*\**p* is significant at <0.01, \*\*\**p* is significant at <0.001

## DISCUSSION

This study serves as the first to demonstrate the feasibility of ULF MRI for paediatric neuroimaging research on antenatal maternal anemia in LMICs, and to corroborate cross-validation work previously reported in healthy adults.^23^ The key finding of this research was evidence for excellent agreement between paired data across HF and ULF systems. Overall, in a pooled sample of infants between 3-18 months of age, there was a very strong linear relationship (Pearsons’s correlations; range of *r =* 0.87 – 0.97) and high agreement (Lins concordance; range of *ρ* = 0.79 – 0.96) between 3T and 64mT brain MRI volume estimates for ICV and ROIs implicated in antenatal maternal anemia. Improved correspondence was observed in older infants, particularly for the basal ganglia structures (caudate nucleus and putamen). While this was also true for ICV and the corpus callosum, findings were less consistent across age groups.

In assessing the overarching validity of ULF MRI for brain volume estimation across different early ages, it is likely that the basal ganglia structures were easier to segment than the corpus callosum due to their high visual contrast. The striatum of the basal ganglia (caudate nucleus and putamen) is a target of the nigrostriatal pathway with dopaminergic neurons projecting from the substantia nigra, resulting in a darker appearance on T2W MRI sequences as indirect consequence of higher iron content.^44,45^ This is particularly prominent for the putamen, which receives the densest dopaminergic input, potentially explaining why this region emerged with the most comparable findings across HF and ULF (*r* = 0.97, *ρ* = 0.96) and perfect agreement between the regression model and line of identity. In contrast, the caudate nucleus has slightly less contrast than the putamen and is closer to the ventricular CSF, making it somewhat more difficult to distinguish structural boundaries for segmentation. This may have contributed to the disagreement observed for this region, although this was marginal with overestimation and underestimation never exceeding 4%. While there was good correspondence across field-strengths for all brain regions investigated here, the results were generally weaker for the corpus callosum with lower concordance values and a regression model that differed significantly from the line of identity. This region may have been more difficult to segment at low field strengths than the distinctive basal ganglia due to it being a narrow and intricate structure of commissural fibers connecting hemispheres, with incomplete myelination in early infancy.^46–48^ Given it’s lower visual contrast on MRI, partial volume effects (PVE), characterized by inaccuracies in voxel representation at ULF strength, may have contributed to segmentation difficulties.

Further investigation revealed general age-related patterns, with improved associations and correspondence between HF and ULF volume estimates in older infants than younger infants. This was demonstrated by a steady increase in associations and concordance from 3 months to 18 months for both the caudate nucleus (*r* = 0.63, *ρ =* 0.61 to *r* = 0.94, *ρ* = 0.92) and putamen (*r* = 0.83, *ρ =* 0.78 to *r* = 0.91, *ρ* = 0.89). This finding is to be expected in the developing brain, as ongoing myelination and maturation contribute to more discrete grey-white matter boundaries, and improved contrast for segmentation of ROIs in children over 12 months of age.^28,49^ While this was generally true for ICV and the corpus callosum with overall improved agreement from 3 months to 18 months, there were inconsistencies across age groups. For example, concordance coefficients for ICV were weakest at 12 months, with ULF 64mT volumes slightly underestimating HF estimates of ICV by 1.77% at 3 months and overestimating them by 0.78% - 4.97% in infants between 6 and 18 months. Similarly, a comparative assessment of the corpus callosum revealed the strongest agreement to be evident at 6 months (*r* = 0.90, *ρ* = 0.79) followed contrastingly by the weakest agreement at 12 months (*r* = 0.82, *ρ* = 0.51). While the ULF 64mT MRI system underestimated corpus callosum HF 3T volumes in infants at 3 months by 5.57%, the extent to which it overestimated HF corpus callosum volumes in older infants was much greater, particularly at 12 (12.15%) and 18 (14.72%) months.

In interpreting this finding, we acknowledge that there was a group difference in age for the 12-month age group, with children being slightly older at the time of their HF 3T scan than their ULF 64mT scan. This may have biased cross-validation comparisons of total brain volume (ICV). Furthermore, given the corpus callosum’s dynamic and non-linear postnatal maturation,^47^ with myelination occurring in a spatiotemporal pattern,^46,48^ it is possible that analyses may have been undermined by the comparison of this ROI at different stages of development. Previous research has demonstrated region-specific developmental trajectories with normal growth spurts occurring in the genu (anterior corpus callosum) at 2-3 months followed by the splenium (posterior corpus callosum) at 4-6 months,^46^ while postnatal myelination begins in the splenium (posterior corpus callosum) at 3-4 months and progresses rostro-caudally to the genu (anterior corpus callosum) at 6-8 months.^46,48^ With such drastic changes occurring in the first year of life in opposite directions, age-differences in paired data at 12 months serve as a likely explanation for generally weaker cross-validation results. This may be exacerbated by segmentation difficulties at ULF for smaller regions of the corpus callosum body, morphologically defined by the Desikan-Killiany atlas^43^ as the mid-posterior, central, and mid-anterior corpus callosum. In particular, the mid-posterior section which corresponds with the isthmus, is known to be the hardest region to segment due to inter-individual variation and its narrow curved shape transitioning between the body and splenium of the corpus callosum.^50^

Study strengths of this research include the corroboration of established clinical strategies for successfully scanning infants in natural sleep across field strengths and the successful use of a novel template-based approach (MiniMORPH)^32^ for specialized processing of paediatric MRI data. In turn, this supported the feasibility of cross-validation work in a relatively large paediatric sample. While future work would benefit from using more closely matched age groups for valid comparisons, it should also focus on improving templates to minimize any potential biases. The ongoing optimization of the MiniMORPH ^32^ pipeline is particularly relevant for the accurate segmentation of intricate corpus callosum regions in infant samples and is likely to be necessary for other small structures of interest such as the hippocampus. To improve generalizability of this processing method for infants, toddlers, and children, we suggest the derivation of templates from larger paired datasets across a broader age range.

ULF MRI is a rapidly growing field with ongoing advances in hardware and software. For example, continuous improvements in sequence development and software versions may further increase comparability across field strengths. Lastly, for this study, correspondence was demonstrated without the prior application of synthetic enhancement using deep learning image quality transfer (IQT). While this reinforces the true validity of the findings without concern of confabulation in super-resolved images where the input does not match the training dataset,^23^ there is great potential for deep learning approaches to improved ULF scan quality at no additional cost.^30,31^ This is an important and relevant consideration for increased scalability of neuroimaging in LMICs.

In conclusion, this study demonstrates high comparability between and HF (3T) and ULF (64mT) volumes for infant brain regions associated with antenatal maternal anemia. These novel findings represent the first paired dataset to be used to validate the use of ULF (64mT) MRI for ongoing paediatric neuroimaging work on antenatal maternal anemia across LMICs. Given that this is a relevant health priority being investigated globally, this work will be key for the meaningful interpretation of neuroimaging research findings on the impact of anemia on child brain development as well the efficacy of ongoing interventions. This includes large-scale research projects across a collaborative international network known as Ultra- Low-Field Neuroimaging In The Young (UNITY)^22^ in an effort to increase neuroimaging accessibility for improved maternal and child health.

## ABBREVIATIONS

CSF: Cerebrospinal Fluid
CUBIC: Cape Universities Body Imaging Centre
GLHT: General Linear Hypothesis Test
HF MRI: High-Field Magnetic Resonance Imaging
HIV: Human Immunodeficiency Virus
ICU: Intensive Care Unit
IQT: Image Quality Transfer
LMIC: Low- and Middle-Income Country
MOU: Midwife Obstetrics Unit
MRI: Magnetic Resonance Imaging
MRR: Multi-Resolution Registration
PVE: Partial Volume Effects
ROI: Region of Interest
SES: Socio-Economic Status
ULF MRI: Ultra- Low-Field Magnetic Resonance Imaging
UNITY: Ultra- Low-Field Neuroimaging In The Young
QC: Quality Check
WHO: World Health Organization

## ARTICLE INFORMATION

### Author Contributions

*Conceptualization:* J.E. Ringshaw, N.J. Bourke, M.R. Zieff, C.J. Wedderburn, C. Casella., J. O’Muircheartaigh, S. Deoni, S.C.R. Williams, K.A. Donald

*Data Curation:* J.E. Ringshaw, N.J. Bourke, M.R. Zieff, L.E. Bradford, C. Casella, J. O’Muircheartaigh

*Formal Analysis:* J.E. Ringshaw

*Funding Acquisition:* J.E. Ringshaw, S.C.R. Williams, K.A. Donald

*Investigation:* L.E. Bradford, S.R. Williams, M. Miles, *Khula South Africa Data Collection

*Methodology:* J.E. Ringshaw, N.J. Bourke, M.R. Zieff, C. Casella, J. O’Muircheartaigh,

S.C.R. Williams, K.A. Donald

*Project Administration:* M.R. Zieff, D. Herr, M. Miles, C. Bennallick

*Resources:* S.C.R. Williams, S. Deoni, K.A. Donald *Software:* N.J. Bourke, C. Casella, J. O’Muircheartaigh *Supervision:* D.J. Stein, S.C.R. Williams, K.A. Donald

*Validation:* J.E. Ringshaw, N.J. Bourke, M.R. Zieff, C.J. Wedderburn, C. Casella, L.E. Bradford, S.R. Williams, J. O’Muircheartaigh, D.J. Stein, D.C. Alexander, D.K. Jones,

S.C.R. Williams, K.A. Donald

*Visualisation:* J.E. Ringshaw, C. Casella

*Writing – Original Draft:* J.E. Ringshaw, K.A. Donald

*Writing – Review and Editing:* All authors.

*Khula South African Data Collection Team: Lauren Davel, Tembeka Mhlakwaphalwa, Bokang Methola, Khanyisa Nkubungu, Candice Knipe, Zamazimba Madi, Nwabisa Mlandu, Ringie Gulwa, Pamela Madikane

## Acknowledgements

We would like to thank all the mothers and infants who participated in the Khula South Africa birth cohort study and made this research possible. We are also sincerely grateful to all the staff at both the Gugulethu Midwife Obstetric Unit as well as the University of Cape Town Neuroscience Institute who were involved in recruitment and data collection.

## Ethical Considerations

The Khula South Africa birth cohort study received ethical approval from the University of Cape Town Human Ethics Research Committee (HREC; 666/2021; 782/2022). All mothers provided written informed consent for cohort participation at Khula enrolment, and for their children to participate in study-specific procedures across timepoints. Given the longitudinal nature of the cohort, study consent is obtained from mothers annually for this cohort.

## Data Availability

Data used for these analyses have been deposited in *ZivaHub Open*, a public repository. This information can be found at the following DOI: https://doi.org/10.25375/uct.29197649.v1

